# Examining the Association between Mitochondrial Genome Variation and Coronary Artery Disease

**DOI:** 10.1101/2022.02.09.22270723

**Authors:** Baiba Vilne, Aniket Sawant, Irina Rudaka

## Abstract

**Background:** Large-scale genome-wide association studies have identified hundreds of single-nucleotide variants (SNVs) significantly associated with coronary artery disease (CAD). However, collectively, these explain <20% of the heritability.

**Hypothesis:** Here, we hypothesize that mitochondrial (MT) SNVs might present one potential source of this “missing heritability”.

**Methods:** We analyzed 265 MT-SNVs in ∼500,000 UK Biobank individuals, exploring two different CAD definitions: a more stringent (myocardial infarction and/or revascularization; HARD=20,405), and a more inclusive (also angina and chronic ischemic heart disease; SOFT=34,782).

**Results:** In HARD cases, the most significant (P<0.05) associations were for m.295C>T (control region) and m.12612A>G (ND5), found more frequently in cases (OR=1.05), potentially related to reduced cardiorespiratory fitness in response to exercise, as well as for m.12372G>A (ND5) and m.11467A>G (ND4), present more frequently in controls (OR=0.97), previously associated with lower ROS production rate. In SOFT cases, four MT-SNVs survived multiple testing correction (at FDR<5%), all potentially conferring increased CAD risk. Of those, m.11251A>G (ND4) and m.15452C>A (CYB) have previously shown significant associations with body height. In line with this, we observed that CAD cases were slightly less physically active and their average body height was ∼2.00 cm lower compared to controls, both traits known to be related to an increased CAD risk. Gene-based tests identified CO2 associated with HARD/SOFT CAD, whereas ND3 and CYB associated with SOFT cases (P<0.05), dysfunction of which has been related to MT oxidative stress, obesity/T2D (CO2), BMI (ND3) and angina/exercise intolerance (CYB). Finally, we observed that macro-haplogroup I was significantly (P<0.05) more frequent in HARD cases vs. controls (3.35% vs. 3.08%), potentially associated with response to exercise.

**Conclusions:** We found only spurious associations between MT genome variation and HARD/SOFT CAD and conclude that more MT-SNV data in even larger study cohorts may be needed to conclusively determine the role of MT-DNA in CAD.

## 1. Introduction

Coronary artery disease (CAD) and its major complication myocardial infarction is the most common cardiovascular disease and the main leading cause of morbidity and mortality worldwide. CAD is posing a huge socio-economic burden to the society and health systems [65] and its prevalence is expected to increase in the coming years [29, 71, 87]. CAD is a multi-factorial disease with complex etiology, considered to be driven by both environment/lifestyle and genetic factors [22, 28, 101]. Over the last 14 years, several large-scale genome-wide association studies and their meta-analysis have identified numerous common genetic variants associated with CAD risk [20, 27, 44, 77, 79, 85, 88, 96, 97, 106] and explored their functional consequences [1, 12, 49–51, 63, 76, 89, 102, 109]. However, collectively, these variants explain only a small proportion (∼20%) of the disease heritability [20, 56]. Genetic variations of the mitochondrial (MT) DNA have remained out of focus for a long time and present an underexplored potential source of the “missing heritability” of several complex traits, including CAD [11, 45, 91].

The human MT-DNA is a maternally-inherited, double-stranded, circular, histone-free “chromosome” of 16,596 base pairs (bp). Each mitochondrion contains 2 to 10 copies of MT-DNA and, depending on the tissue energy requirement, each human cell may contain hundreds of mitochondria [4]. MT-DNA encodes 37 genes corresponding to subunits ND1 to 6 (and 4L) of respiratory complex I, catalytic subunits I-III (CO1-3) of cytochrome c oxidase (respiratory complex IV), subunits adenosine triphosphate 6 and 8 (ATP6 and 8) of F1F0 ATPase and cytochrome b of respiratory complex III. The remaining genes encode 2 ribosomal RNAs (16S and 12S rRNAs) and 22 transfer RNAs (tRNAs), used for mitochondrial protein synthesis [11, 92, 104]. All of them are involved in oxidative phosphorylation (OXPHOS), the process by which ATP, the major source of energy, is being synthesized [25, 37, 74].

A toxic by-product of OXPHOS is the production of reactive oxygen species (ROS), unstable compounds which can generate free radicals [58]. Mitochondria are the primary source of endogenous ROS [58]. By antioxidant defense, cells can manage certain levels of free radical production. However, if threshold levels are exceeded, a state of oxidative stress occurs [68], which is known to play a vital role in the pathogenesis of atherosclerosis and CAD [69, 102]. Many of the common CAD risk factors such as age, hypertension, hyperglycemia, high cholesterol levels and smoking are also known to perturb mitochondrial function and increase oxidative stress [11]. In particular, the lifestyle of modern societies is increasingly related to reduced physical activity and calorically excessive diets [21, 70], favoring ROS production and possibly increasing the risk of CAD [61, 82], due to mitochondrial dysfunction, reduced bioenergetic capacity and disrupted redox homeostasis [6, 67].

Although the role for mitochondrial dysfunction in CAD etiology is well-established, the role of the mitochondrial genome in this process has not been extensively investigated [11]. Several forms of cardiovascular disease have been related to the presence of pathogenic mitochondrial genome mutations. However, the vast majority of mitochondrial genetic variation, “natural” single nucleotide variants (SNVs) have not been directly linked to disease pathogenesis [11]. During evolution, a number of such MT-SNVs have accumulated in mitochondrial genomes subdividing the human population into several discrete (geographic region specific) mitochondrial phylogenetic clades or haplogroups [95]. As the mitochondrial genome does not undergo DNA recombination, haplogroups are relatively stable and enable the clustering of individuals based on their shared maternal ancestry [95]. These clusters are often associated with different racial/ethnic groups [11]. Considering that family history and race/ethnicity is known to influence CAD risk, it is reasonable to assume that mitochondrial haplogroups may contribute to this heritable modulator of CAD susceptibility [11].

Increasing evidence suggests that non-pathogenic “natural” MT-SNVs and haplogroups may be associated with alterations in mitochondrial function [11, 25, 103], including subtle differences in OXPHOS capacity and the generation of ROS [30, 84], thus, potentially influencing the onset of and differences in CAD risk of different human populations [11, 80]. For example, MT-SNVs m.8701A>G and m.10398A>G have been demonstrated to lead to lower matrix pH and alterations in intracellular calcium levels [48]. In addition, several other MT-SNVs have been previously identified as significantly associated with HDL cholesterol and triglycerides levels [34], as well as with several metabolic traits including insulin levels and body weight [57]. Mitochondria bearing macro-haplogroup H-defining SNVs have been observed to possess several differences in basal oxygen consumption, maximal respiratory capacity and oxidant production relative to other European haplogroups, especially macro-haplogroup J [72] and African macro-haplogroup L [59]. Similarly, macro-haplogroup T has been associated with CAD and diabetic retinopathy in Middle European Caucasian populations [55]. On the other hand, several haplogroups in both Europe and Asia have been related to increased longevity, likely via reduced oxidant generation, which is a driver of the ageing process [16, 23].

In this study, we hypothesize that mitochondrial genome variation might present a potential source of the “missing heritability” of CAD. To explore this hypothesis, we performed: (1) association analyses of common/low-frequency MT-SNVs (MAF>0.01; N=111) with CAD; (2) gene-based tests to investigate the cumulative impact of all MT-SNVs on the mitochondrial genes in relation with CAD and (3) comparisons of mitochondrial haplogroup frequencies of individuals with CAD. In all cases, we explored two different CAD definitions (as previously used by Nelson et al. [77]): a more stringent (HARD=20,405), considering only myocardial infarction and/or revascularization and a more inclusive (SOFT=34,782), including all HARD CAD cases, as well as also angina and chronic ischemic heart disease vs. controls in a cohort of ca. 500,000 UK Biobank individuals. The complete workflow of this analysis is summarized in Fig.1.

**Figure 1.**
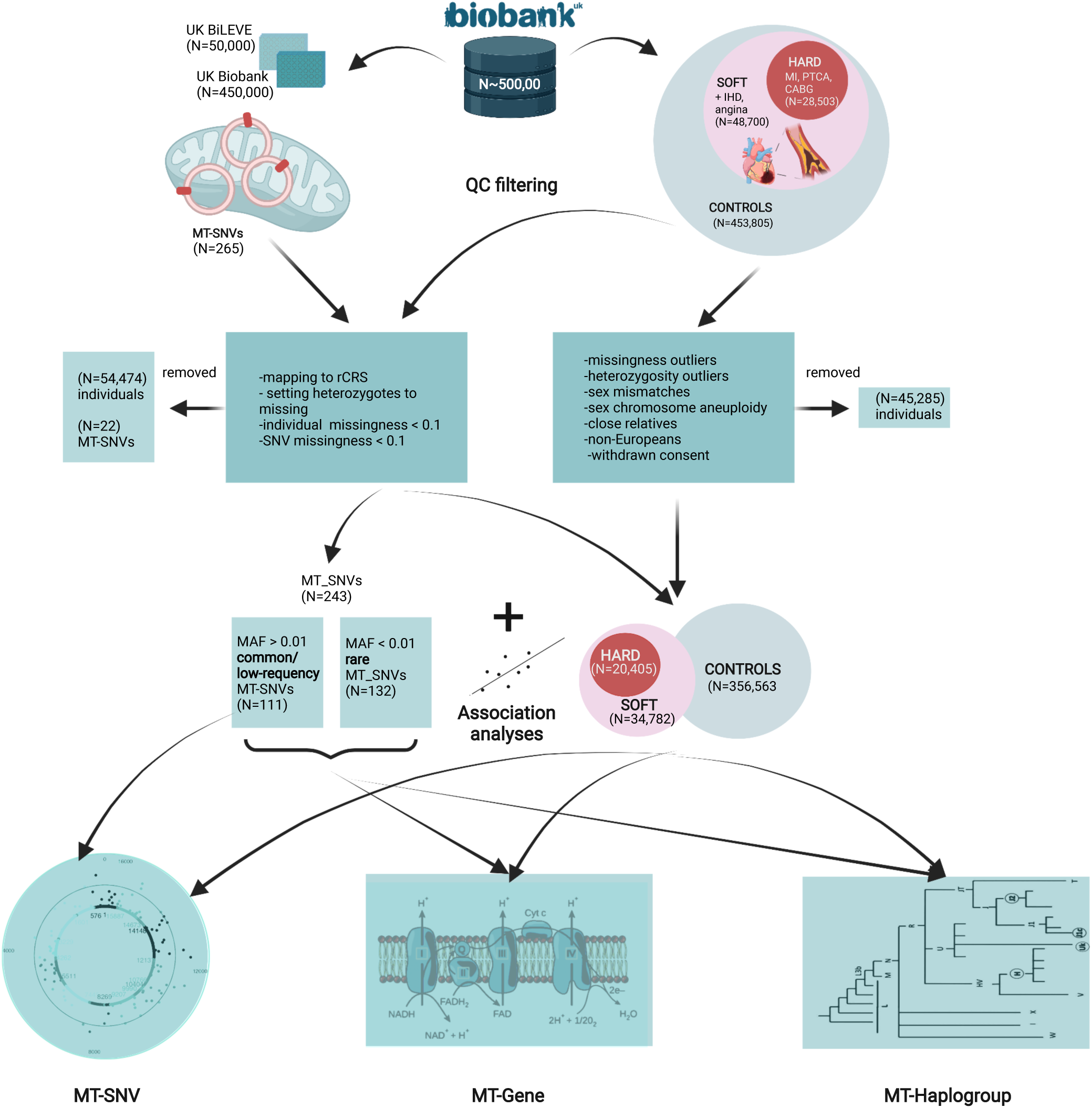
A complete workflow of the analyses performed.

## 2. Materials and Methods

### 2.1. Study population, disease phenotypes and quality filtering

The UK Biobank [13] is a large population-based prospective cohort study from the United Kingdom with genetic and deep phenotypic (∼7,221 phenotypes http://www.nealelab.is/uk-biobank) data on ca. 500,000 individuals aged 40 to 69. We downloaded these data (application ID 61684) and used similar CAD case definition, as previously described by Nelson et al. [77] for UK Biobank. HARD CAD cases included individuals with fatal or non-fatal myocardial infarction (MI), percutaneous transluminal coronary angioplasty (PTCA) or coronary artery bypass grafting (CABG). SOFT CAD included individuals meeting the HARD CAD definition as well as those with chronic ischemic heart disease (IHD) and angina (Fig.1). In HESIN hospital episodes’ data and death registry data from diagnosis and operation (primary and secondary causes), MI was defined as hospital admission or cause of death due to ICD9 410-412, ICD10 I21-I24, I25.2; PTCA was defined as hospital admission for PTCA (OPCS-4 K49, K50.1, K75); CABG was defined as hospital admission for CABG (OPCS-4 K40-K46); and angina or chronic IHD was defined as hospital admission or death due to ICD9 413, 414.0, 414.8, 414.9, ICD10 I20, I25.1, I25.5-I25.9. In UK Biobank self-reported data, cases were defined as having ‘vascular/heart problems diagnosed by doctor’ or ‘non-cancer illnesses that self-reported as angina or heart attack’. Self-reported surgery included PTCA, CABG or triple heart bypass. All participants not defined as CAD cases using the SOFT definition were considered as controls in the analysis. For a complete list of definition codes, see Supplementary Table 1. We subsequently performed individual-level filtering (Fig.1) by removing missingness or heterozygosity outliers, participants with self-reported vs. genetically inferred sex mismatches or putative sex chromosome aneuploidy, individuals that were not of European (EUR) ancestry and individuals having withdrawn their consent at the time of analysis. We also identified closely related participants (kinship coefficient >0.088 i.e., first or second-degree relative pairs), preferentially retaining CAD cases or relative with the highest call-rate.

The following individual characteristics were also extracted in order to characterize HARD/SOFT CAD cases vs. controls: age at recruitment (field #21022), sex (field #31), BMI (field #21001), height (field #50), hypertension (fields #4080 and #4079), hypercholesterolemia (self-reported data and ICD9/10) and (self-reported) use of cholesterol lowering drugs, insulin and blood pressure medications (field #6153), type 2 diabetes (T2D, fields #41201, #41202 and #4120, E11), glycemic control, obesity, smoking status (‘ever smoked’: field #20160 and ‘current’ from ‘smoking status’ field #20116), family history of heart disease (fields #20107, #20110, #20111 in any first-degree relative, i.e. father, mother or sibling, respectively). Data at the time of first assessment were obtained and processed to binary (yes / no) values or mean values for fields with continuous data with multiple readings at the time of first assessment. The statistical tests used were Mann-Whitney test for comparison of the continuous measures between HARD vs. SOFT cases vs. controls and the Chi-square test for goodness of fit, when comparing nominal/binary data in the same groups.

### 2.2. Genotype data quality control

In UK Biobank [13], genotyping was performed using Affymetrix UK biobank Axiom (450,000 samples) and Affymetrix UK BiLEVE Axiom (50,000 samples) arrays (Fig.1) and the autosomal genetic data were then imputed to the Haplotype Reference Consortium panel and UK10K4 + 1000 Genomes panel. We downloaded the genotype data for the 265 MT DNA variants for all 500,000 individuals and pre-processed MT DNA data as previously described in ref. [57]. In brief, we first made sure that the reference alleles match the latest MT Cambridge Revised Sequence (rCRS) of the Human MT DNA positions. After setting all potential heterozygotes to missing, further quality control of genotyped individuals included filtering for missingness by individual <0.1 and missingness by SNV<0.1 with PLINK [14]. For common/low-frequency variant association analyses, we also required that the minor allele frequency (MAF)>0.01. An overview of the filtering of MT-SNVs is provided in Fig.1

### 2.3. MT-SNV association analyses

For common and low-frequency (MAF >0.01; n=111) variants, we performed single marker tests to explore their associations with HARD and SOFT CAD (Fig.1) using SNPTEST v2.5.4 with the frequentist test and expected method, as previously described by ref. [9]. We used as covariates the array (UK Biobank vs UK BiLEVE), sex, birth year and the first five principal components of the autosomal genotype data, provided by the UK Biobank, similar to ref. [77] and Benjamini-Hochberg (BH) [8] adjustment for multiple testing was applied to calculate the false discovery rate (FDR). MT-SNV annotations were performed using the manually-curated database, HmtVar (https://www.hmtvar.uniba.it/).

### 2.4. MT-gene-based association analyses

To also consider the potential effects of rare (MAF ≤ 0.01) variants on CAD risk, we assigned all SNVs to MT genes based on MITOMAP https://www.mitomap.org/MITOMAP and used the R software package SKAT (v2.0.1) [62] to perform MT-gene-based (additionally including the whole mitochondrion as our region of interest, MT) association analyses with HARD and SOFT CAD phenotypes (Fig.1), again using as covariates the array (UK Biobank vs UK BiLEVE), sex, birth year and the first five principal components of the autosomal genotype data, provided by the UK Biobank, similar to ref. [77] and obtain resampled residuals (n.Resampling=1000, type.Resampling=“bootstrap”) to compute resampling P value.

### 2.5. Haplogroup assignment

We used the PhyloTree Build 17 [98] as implemented in HaploGrep (v2.2.8) [53] to estimate mitochondrial haplogroups in our dataset. Thereafter, we assigned individuals to one of the major European haplogroups (H, I, J, K, R, T, U, V, W, X), or to a group of ‘others’ https://www.mitomap.org/foswiki/pub/MITOMAP/WebHome/simple-tree-mitomap-2019.pdf. Fisher’s exact test [31] was used to calculate the statistical significance of the overlaps between haplogroups and HARD and SOFT CAD phenotypes. Benjamini-Hochberg (BH) [8] adjustment for multiple testing was applied to calculate the false discovery rate (FDR) (Fig.1).

## 3. Results

### 3.1. Characteristics of study subjects

The current study included ca. 500,000 genotyped individuals from the UK Biobank [13] 48,700 with an inclusive CAD phenotype (SOFT) that incorporated self-reported angina or other evidence of chronic coronary heart disease, of which 28,503 had a more stringently defined CAD phenotype (HARD) of myocardial infarction (Fig.1), similarly as in ref. [77]. All participants (N=453,805) not defined as CAD cases using the SOFT definition will be considered as controls in the analysis. After this step of quality control 45,285 individuals were removed, 24,770 cases with the HARD, 42,079 cases with the SOFT CAD phenotype and 415,271 controls remained. Finally, further quality control of genotyped individuals included filtering for missingness by individual <0.1 and missingness by SNV<0.1 was performed, considering the 265 mitochondrial variants present on the UK Biobank or UK BiLEVE arrays (as described in section 2.2. in Methods). As a result, a further set of 54,474 individuals were removed, leaving us with 20,405 cases with the HARD and 34,782 cases with the SOFT phenotype vs. 356,563 controls. Individual characteristics, in terms of common CAD risk factors, of these individuals are summarized in Table 1.

**Table 1.** Individual characteristics of HARD and SOFT CAD cases vs. controls. *** Represents statistically significant (of P<0.001) difference between HARD and SOFT CAD cases vs. controls, whereas +++ represents statistically significant (of P<0.001) difference between HARD vs. SOFT CAD cases.

| Risk factors | HARD*** | SOFT*** | CONTROL |
| --- | --- | --- | --- |
| Men (%) | 77 | 67 | 43 |
| Age, years (mean±SD, range) | 63 (61.33±6.35, 58-66) | 63 (61.14±6.43, 58-66) | 57 (56.08±8.05, 50-63) |
| Diastolic blood pressure >90 mmHg (%) | 21.23 | 21.54 | 22.41 |
| Systolic blood pressure >140 mmHg (%) | 49.88 | 50.04 | 41.52 |
| Hypercholesterolemia (%) | 51.20 | 44.78 | 6.00 |
| Hypertriglyceridemia (%) | 1.55 | 1.43 | 0.80 |
| Poor glycemic control (%) | 3.59 | 3.20 | 0.80 |
| Type 2 Diabetes (%) | 20.80 | 19.41 | 4.14 |
| BMI, kg/m2 (mean±SD, range) | 28.29 (28.91± 4.72, 25.72-31.47) | 28.34 (29.03±4.98, 25.66-31.66) | 26.51 (27.18±4.69, 23.96-29.60) |
| Obesity (BMI > 30 kg/m2, %) | 35.17 | 36.10 | 22.65 |
| Central obesity (%) | 63.39<sup>+++</sup> | 60.40 | 36.61 |
| Body height <u>Male</u> | 173.92±6.76<sup>+++</sup> (Med=174) | 174.16±6.82 (Med=174) | 175.95±6.82 (Med=176) |
| (mean±SD, range): Female | 160.37±6.38<sup>++</sup> (Med=160) | 160.68±6.33 (Med=161) | 162.61±6.29 (Med=163) |
| Physically active (%) | 51.31 | 51.08 | 54.60 |
| Smoking history (ever smoked, %) | 72.23<sup>+++</sup> | 69.56 | 60.21 |
| Current Smoker (%) | 13.74<sup>+</sup> | 12.90 | 9.27 |
| History of Heart Disease in First Degree Relative(%) | 59.14<sup>+++</sup> | 57.35 | 41.41 |

On average, 77/67% vs. 43% of the individuals were men in HARD/SOFT CAD cases vs. controls, respectively and the individuals’ mean age was 63 years in cases vs. 57 years in controls. 49.88%/50.04% vs. 41.52% of the individuals displayed Systolic blood pressure >140 mmHg. Hypercholesterolemia was reported by 51.20%/44.78% vs. 6.00% HARD/SOFT CAD cases vs. controls, respectively, while hypertriglyceridemia and poor glycemic control (defined as serum triglyceride levels >5.6 mmol/dL and HbA1c levels >64 mmol/mol, respectively) was found in 1.55%/1.43% and 3.59%/3.20% HARD/SOFT CAD cases vs. 0.80% (both traits) in controls, respectively. 20.80%, 19.41% and 4.14% had T2D. On average, the BMI of cases was 28.00 kg/m2 vs. 26.51 kg/m2 in controls and 35.17%/36.10% HARD/SOFT CAD cases vs. 22.65% controls displayed obesity (defined as BMI ≥ 30 kg/m2). The average body height of HARD/SOFT CAD cases was 173.92/174.16 cm (male) and 160.37/160.68 cm (female) vs. 175.95 cm and 162.61 cm in male and female controls, respectively. Sufficient physical activity (i.e., meeting the 2017 UK Physical activity guidelines of 150 minutes of moderate activity per week or 75 minutes of vigorous activity) was seen in 51.31%/51.08% and 54.60% of HARD/SOFT CAD cases vs. controls, respectively. 72.23%/69.56% vs. 60.21% of the individuals had a positive smoking history (answered ‘yes’ to ‘ever smoked’) and 13.74%/12.90% vs. 9.27% were current smokers. Finally, 59.14%, 57.35% and 41.41% had positive family history of heart disease. In all cases, the differences between CAD cases and controls were statistically significant (P<0.001, Table 1).

### 3.2. MT-SNV associations with HARD and SOFT CAD phenotypes

After quality control of genotyped individuals (including filtering for missingness by individual <0.1 and missingness by SNV<0.1, as described in section 2.2. in Methods), from the 265 MT-SNVs present on the UK Biobank or UK BiLEVE arrays, 243 remained for further analyses. For the genotyped common and low-frequency MT-SNVs (MAF>0.01; N=111, of those N=39 with MAF>0.05; genotyping rate >0.99) in UK Biobank, we performed single marker association analyses with HARD (N=20,405) and SOFT (N=34,782) CAD phenotypes, adjusting for the array, sex, birth year and first five principal components.

In HARD cases, no MT-SNVs survived multiple testing correction, the most significant (nominal P<0.05) findings (Table 2 and Fig.2) being for m.295C>T (rs41528348, P=0.0118, MAF=0.10, OR=1.05; 95% CI 1.02-1.09, in control region/CR, tagging macro-haplogroup J) and m.12612A>G (rs28359172, P=0.0158, MAF=0.10, OR=1.05; 95% CI 1.02-1.08, synonymous, in ND5 gene, tagging macro-haplogroup J), both more frequent in cases, thus potentially conferring increased CAD risk. In addition, four more MT-SNVs were found more frequently in controls: m.12372G>A (rs2853499, P=0.0059, MAF=0.22, OR=0.97; 95% CI 0.95-0.99, synonymous, in ND5 gene, tagging macro-haplogroup U), m.11467A>G (rs2853493, P=0.0065, MAF=0.22, OR=0.97; 95% CI 0.95-1.00, synonymous, in ND4 gene, tagging macro-haplogroup U), m.15301G>A (rs193302991, P=0.0115, MAF=0.04, OR=0.97; 95% CI 0.92-1.03, synonymous, in CYB gene) and m.7768A>G (rs41534044, P=0.0185, MAF=0.04, OR=0.91; 95% CI 0.86-0.96, synonymous, in CO2 gene). For a complete list of MT-SNV associations with HARD CAD phenotypes, see Supplementary Table 2.

**Figure 2.**
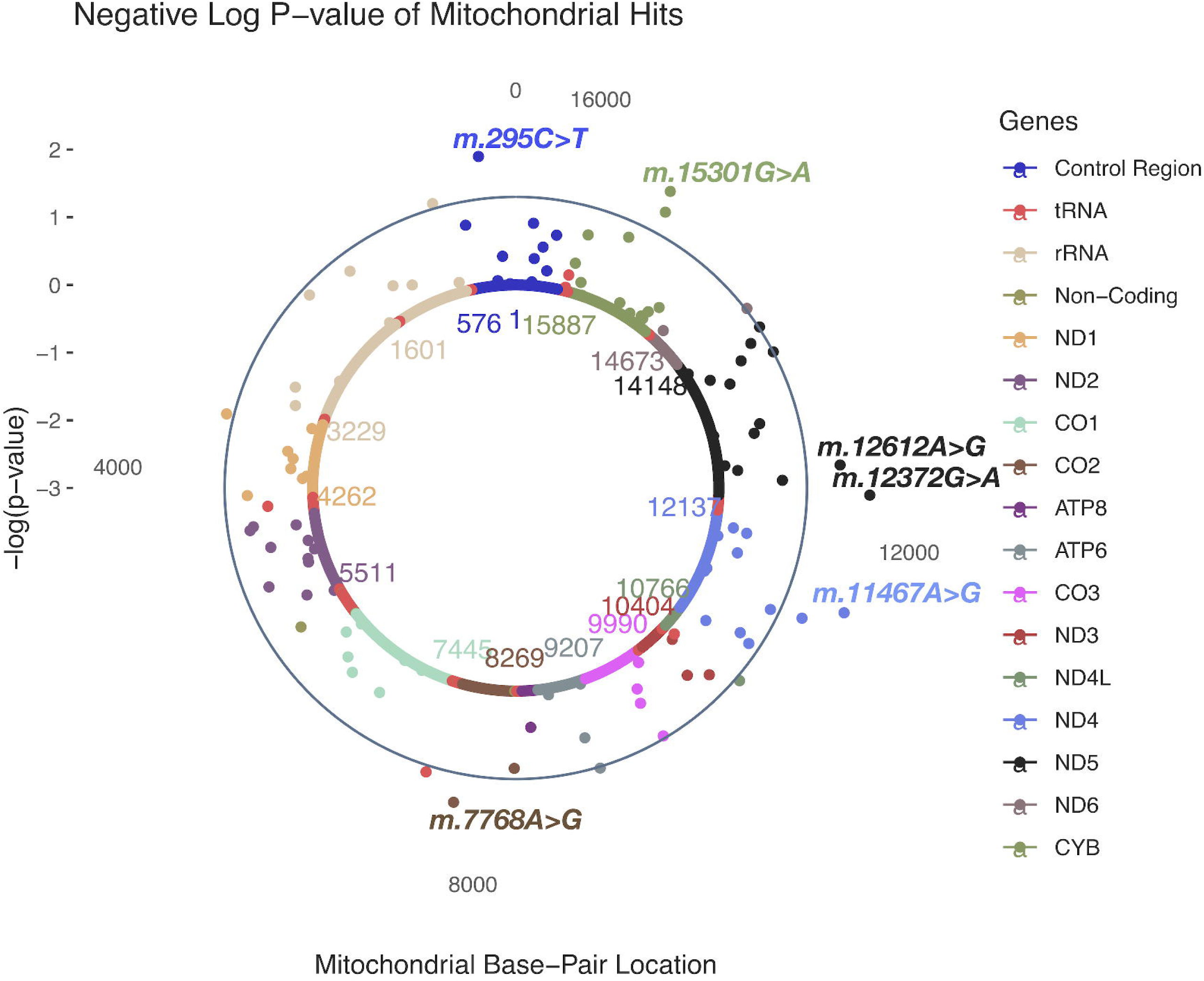
A solar plot of HARD CAD and common and low-frequency (MAF>0.01; N=111) MT SNV associations.

**Table 2.** HARD CAD and common and low-frequency (MAF>0.01; N=111) MT SNV most significant associations. CR=control region.

| Locus | RSID | Variation | MAF | AA | OR | 95% CI | P | HG |
| --- | --- | --- | --- | --- | --- | --- | --- | --- |
| *ND5* | rs2853499 | m.12372G>A | 0.22 | Syn | 0.97 | 0.95-0.99 | 0.0059 | U |
| *ND4* | rs2853493 | m.11467A>G | 0.22 | Syn | 0.97 | 0.95-1.00 | 0.0065 | U |
| *CYB* | rs193302991 | m.15301G>A | 0.04 | Syn | 0.97 | 0.92-1.03 | 0.0115 | . |
| *CR* | rs41528348 | m.295C>T | 0.10 | . | 1.05 | 1.02-1.09 | 0.0118 | J |
| *ND5* | rs28359172 | m.12612A>G | 0.10 | Syn | 1.05 | 1.02-1.08 | 0.0158 | J |
| *CO2* | rs41534044 | m.7768A>G | 0.04 | Syn | 0.91 | 0.86-0.96 | 0.0185 | . |
| *ND4* | rs28358285 | m.11299T>C | 0.08 | Syn | 0.94 | 0.91-0.98 | 0.0227 | K |
| *CYB* | rs41518645 | m.15257G>A | 0.02 | Asp → Asn | 1.11 | 1.04-1.19 | 0.0231 | . |
| *ND1* | rs28358584 | m.3480A>G | 0.08 | Syn | 0.94 | 0.91-0.98 | 0.0390 | . |
| *tRNA<sup>Ser(UCN)</sup>* | rs201950015 | m.7476C>T | 0.02 | . | 1.01 | 1.03-1.19 | 0.0400 | . |
| *rRNA<sup>12S</sup>* | rs2853518 | m.750A>G | 0.02 | . | 0.87 | 0.80-0.95 | 0.0420 | . |
| *ND4L* | rs28358280 | m.10550A>G | 0.08 | Syn | 0.95 | 0.91-0.98 | 0.0435 | K |
| *ND6* | rs193302977 | m.14167C>T | 0.08 | Syn | 0.95 | 0.91-0.98 | 0.0476 | . |
| *ATP6* | rs193303045 | m.9055G>A | 0.09 | Ala → Thr | 0.95 | 0.92-0.98 | 0.0476 | . |
| *ND5* | rs869156190 | m.13965T>C | 0.01 | Syn | 1.10 | 1.00-1.21 | 0.0484 | . |
| *ND5* | rs28359178 | m.13708G>A | 0.12 | Ala → Thr | 1.04 | 1.01-1.07 | 0.0499 | J |

In SOFT cases, four MT-SNVs survived multiple testing correction (at FDR<5%; Table 3 and Fig.3), all potentially conferring increased CAD risk: m.10400C>T (rs28358278, P=0.0007, MAF=0.02, OR=1.28; 95% CI 1.21-1.35, non-synonymous/Thr → Ala, in ND3 gene, tagging macro-haplogroup M), m.11251A>G (rs869096886, P=0.0011, MAF=0.20, OR=1.03; 95% CI 1.01-1.05, synonymous, in ND4 gene, tagging macro-haplogroups J and T), and two MT-SNVs in CYB gene - m.15452C>A (rs193302994, P=0.0017, MAF=0.20, OR=1.03; 95% CI 1.01-1.05, non-synonymous/Leu → Ile, tagging macro-haplogroups J and T) and m.15301G>A (rs193302991, P=0.0010, MAF=0.04, OR=1.03; 95% CI 0.99-1.07, synonymous). For a complete list of MT-SNVs associations with SOFT CAD phenotype, see Supplementary Table 3.

**Figure 3.**
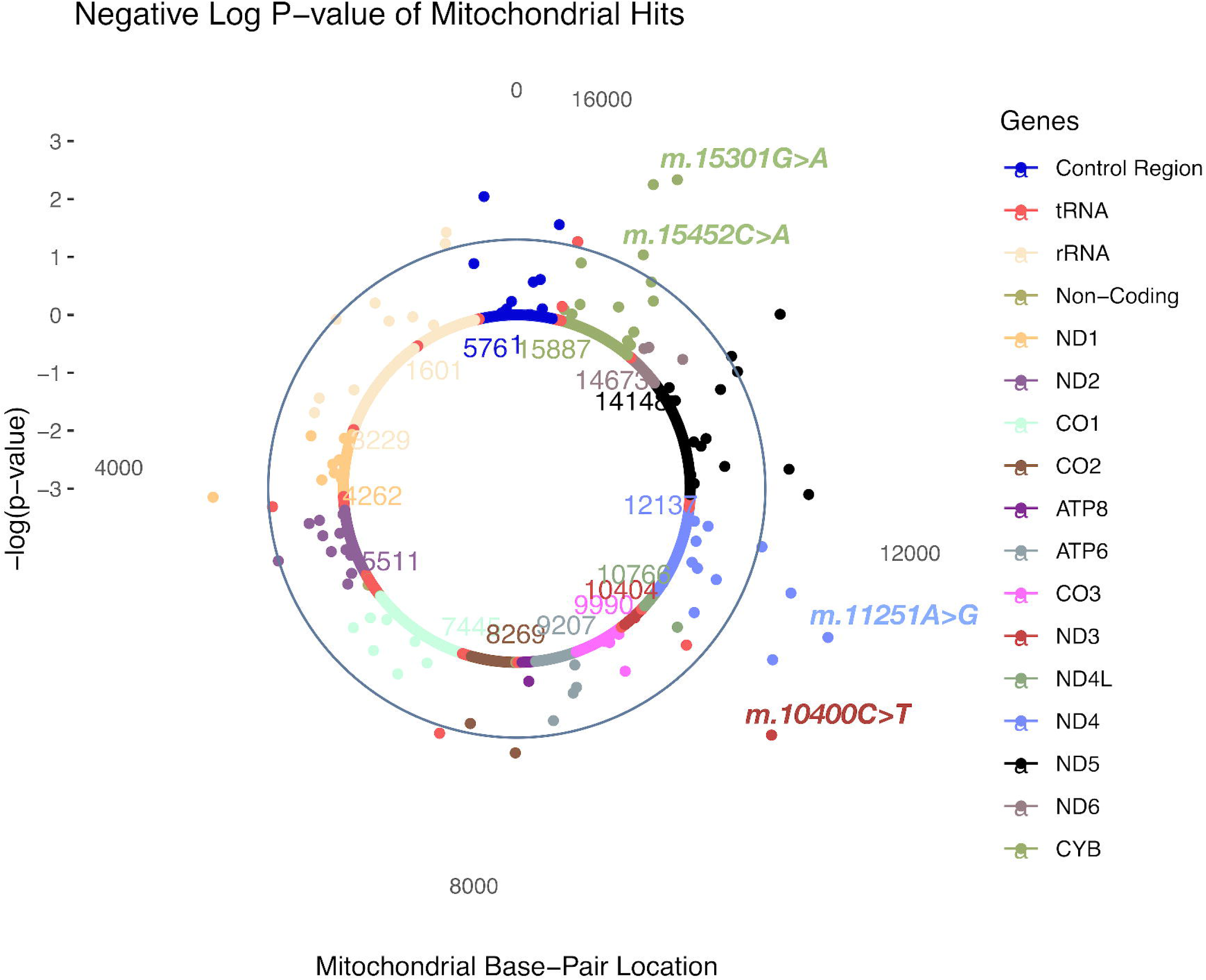
A solar plot of SOFT CAD and common and low-frequency (MAF >0.01; N=111) MT SNV associations. CR=control region.

**Table 3.**
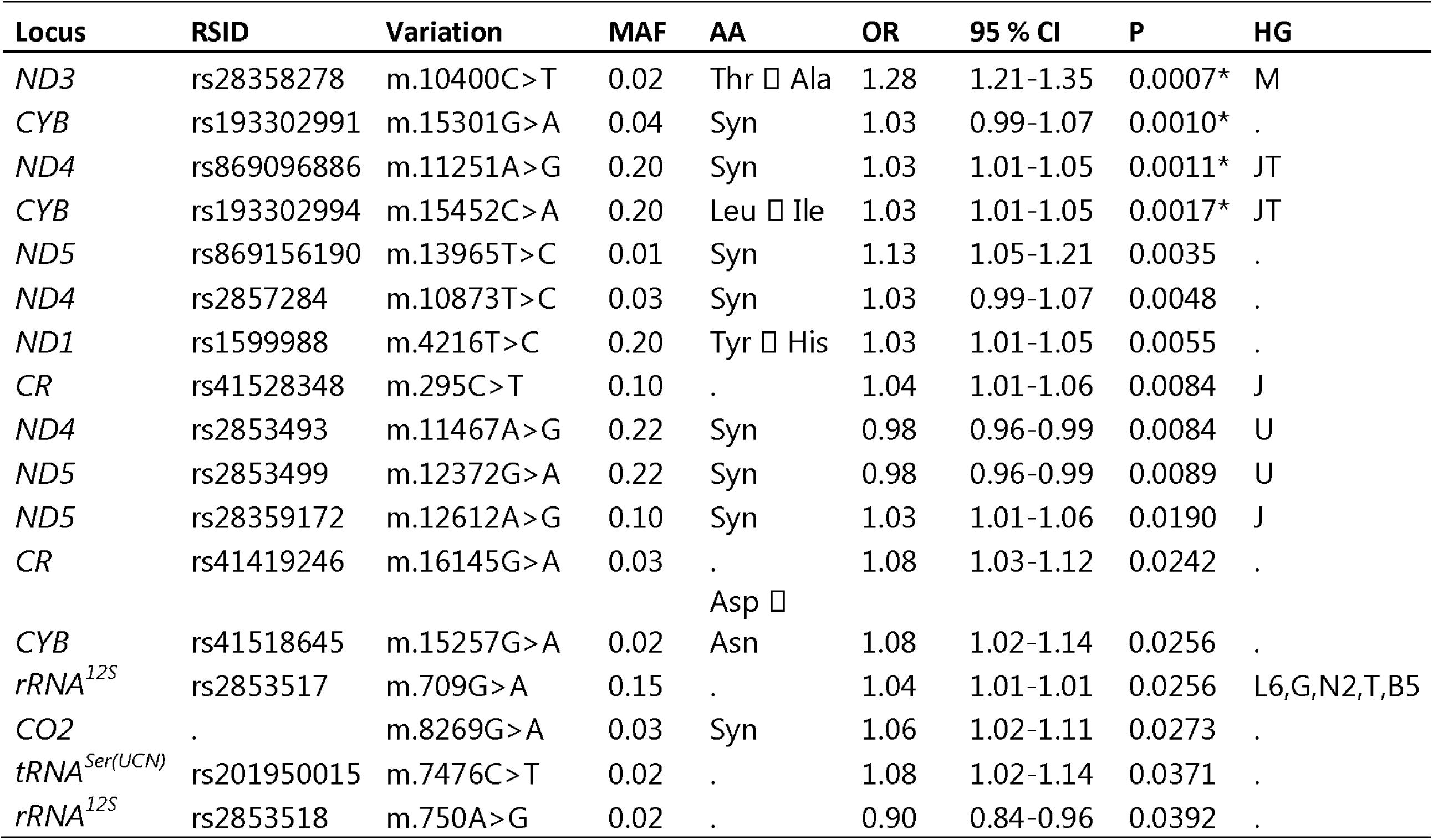
SOFT CAD and common and low-frequency (MAF>0.01; N=111) MT SNV most significant associations. CR=control region. *MT-SNVs that survived multiple testing correction (at FDR<5%).

### 3.3. MT gene-based associations with HARD and SOFT CAD phenotypes

We next sought to also consider the potential effects of rare (MAF ≤0.01) MT-SNVs on CAD risk, hence we performed MT-gene-based association analyses with HARD and SOFT CAD phenotypes, additionally including the whole mitochondrion as our region of interest (MT). As a result, we observed that in both, HARD and SOFT cases, CO2 displayed gene-based association at nominal significance (P<0.05), while CYB and ND3 were also associated (nominal P<0.05) with SOFT CAD phenotype (Table 4). When considering the whole mitochondrion (MT), no significant associations with CAD were observed (N=243; P=0.07, Table 4).

**Table 4.**
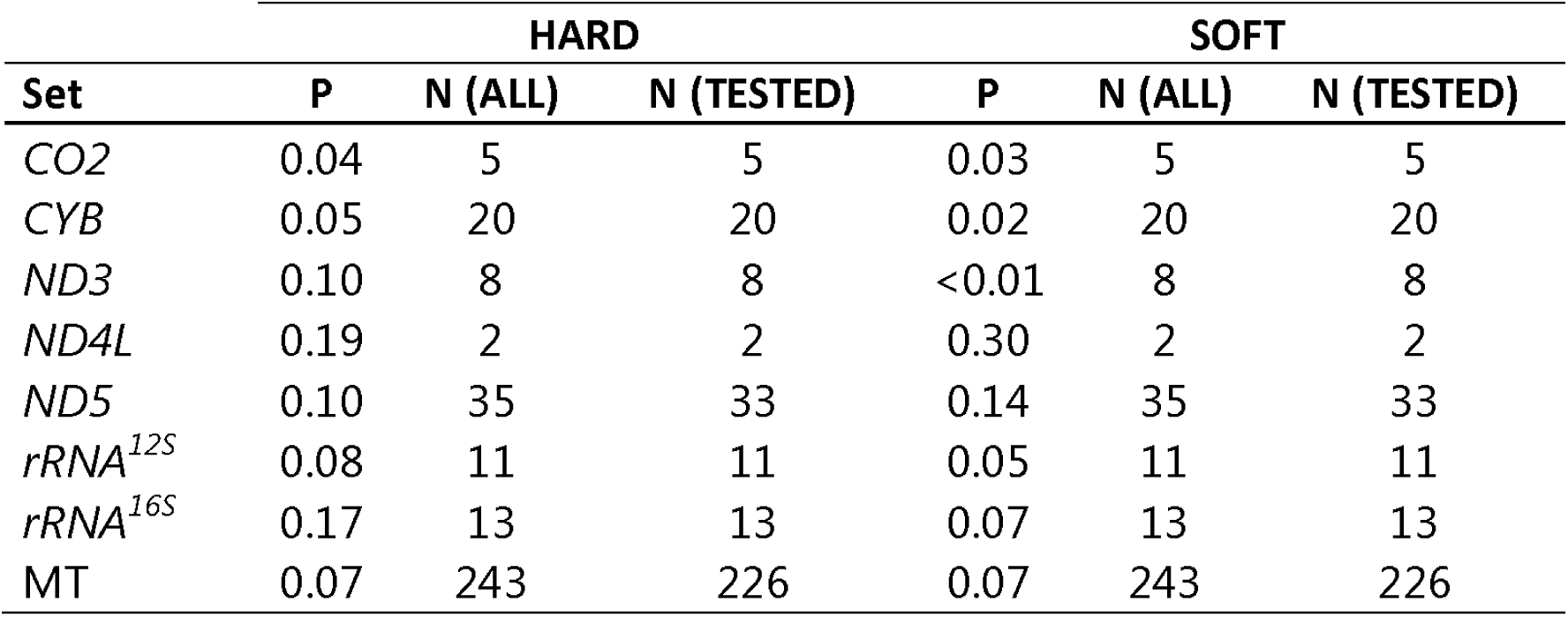
MT gene-based associations with HARD and SOFT CAD phenotypes. MT=mitochondrion; N (ALL)=the number of MT-SNVs in the region; N (TESTED)=the number of MT-SNVs from the region considered in the gene-based test.

### 3.4. MT-haplogroup associations with HARD and SOFT CAD phenotypes

Different human mitochondrial haplogroups may result in differences in mitochondrial function that may influence susceptibility to CAD. Hence, we estimated all the mitochondrial haplogroups in our dataset (Table 5 and Supplementary Tables 4 and 5).

**Table 5.**
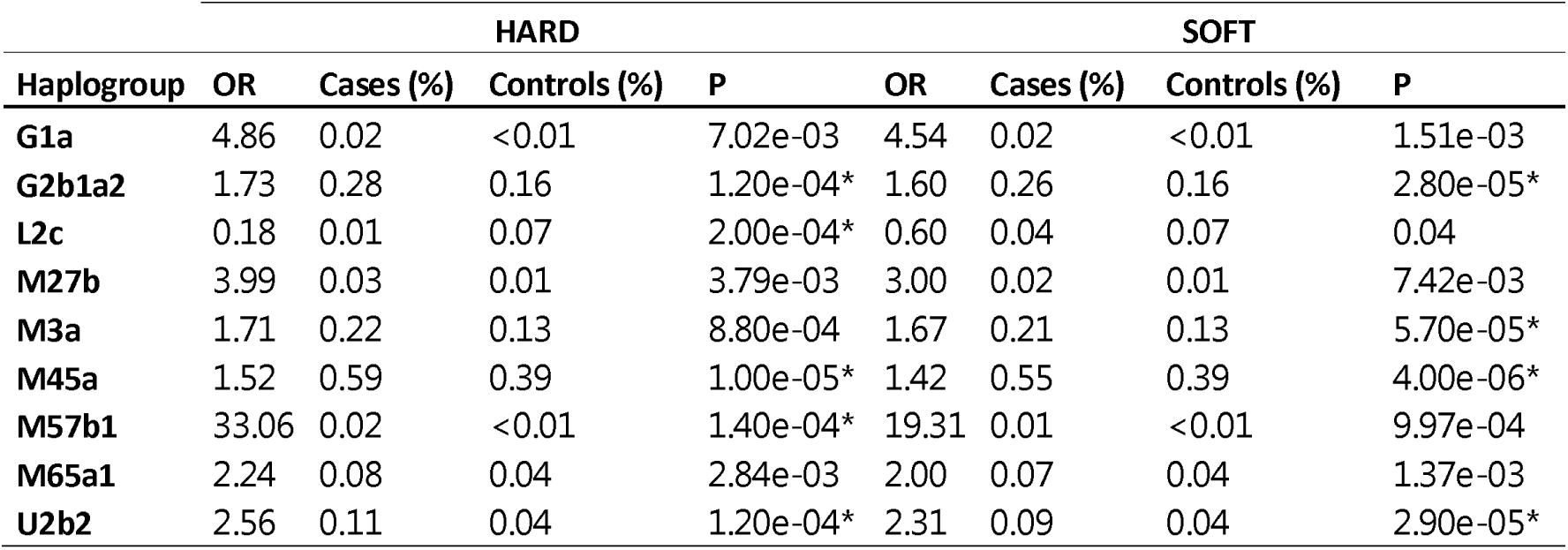
Haplogroup assignment in HARD and SOFT CAD cases vs. controls prior to further assigning individuals to one of the major European haplogroups. * Indicates that a haplogorup survived multiple testing correction (at FDR<5%).

Three haplogroups survived multiple testing correction (at FDR<5%) in both HARD and SOFT cases vs. controls: M45a (0.59% and 0.55 % vs. 0.39%, OR=1.52 and OR=1.42, respectively), G2b1a2 (0.28% and 0.26% vs. 0.16%, OR=1.73 and OR=1.60, respectively) and U2b2 (0.11% and 0.09% vs. 0.04%, OR=2.56 and OR=2.31, respectively). In HARD cases, also haplogroup M57b1 was significantly (at FDR<5%) over-represented in cases vs. controls (0.02% vs. <0.01%, OR=33.06), while haplogroup L2c was significantly (at FDR<5%) under-represented cases vs. controls (0.01% vs.0.07%, OR=0.18) (Table 5 and Supplementary Table 4). In SOFT cases, also haplogroup M3a was significantly (at FDR<5%) over-represented in cases vs. controls (0.21% vs. 0.13%, OR=1.67, Table 5 and Supplementary Table 5).

When further assigning individuals to one of the major European haplogroups (Fig.4). As a result, 43.28%, 3.19%, 10.70%, 8.25%, 0.22%, 9.52%, 13.70%, 2.65%, 2.01%, 1.34% and 5.14% of individuals belonged do the haplogroup H, I, J, K, R, T, U, V, W, X or ‘Others’, respectively. Overall, the frequencies of the major European mitochondrial haplogroups did not differ significantly (at FDR<5%) between CAD patients and control subjects (Fig.4). Only the frequency of haplogroup I was significantly (nominal P<0.05) higher in patients with HARD CAD phenotype vs. controls (3.35% vs. 3.08%, OR=1.09) and the haplogroup R was significantly (nominal P<0.001 and P<0.01) higher in patients with HARD and SOFT CAD phenotype vs. controls (0.26% and 0.23% vs. 0.16%, OR=1.70 and OR=1.49, respectively; Fig.4).

**Figure 4.**
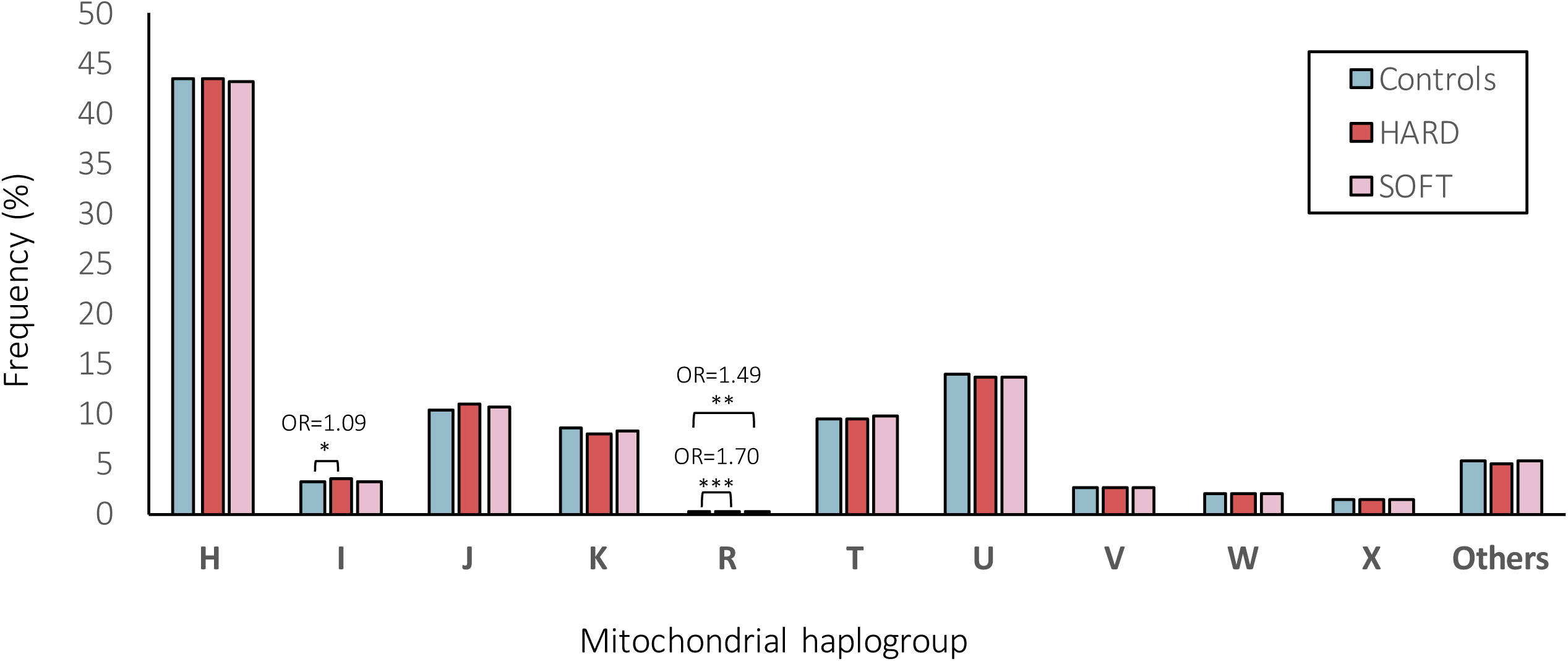
Frequencies (%) of mitochondrial (MT) haplogroups within HARD and SOFT CAD phenotypes vs. controls.

## 4. Discussion

Over the last 14 years, several large-scale genome-wide association studies have found hundreds of single-nucleotide variants (SNVs) significantly associated with CAD, however, these explain <20% of the heritability. In this study, we hypothesize that mitochondrial (MT) SNVs might present one potential source of the “missing heritability”.

We analyzed 265 common/low-frequency (MAF ≥1%) and rare (MAF<1%) MT-SNVs in ∼500,000 UK Biobank individuals, exploring two different CAD definitions, HARD (N=20,405) and SOFT (N=34,782) (Fig.1.), as previously proposed by [77], and using the array, sex, birth year and first five principal components as covariates. Overall, the differences in the prevalence of common risk factors among CAD cases (both HARD and SOFT phenotypes) and controls were statistically significant (P<0.001; Table 1), male gender, older age, hypertension, hypercholesterolemia, obesity, T2D, physical inactivity, shorter body statue, smoking and positive family history demonstrating predominance in CAD patients.

When performing common and low-frequency MT SNVs (MAF>0.01; N=111) association analyses in these individuals, we observed that in HARD cases, no MT-SNVs survived multiple testing correction, the most significant (nominal P<0.05) findings being for m.295C>T, m.12612A>G, m.12372G>A, m.11467A>G, m.15301G>A and m.7768A>G (Table 2 and Fig.2). m.295C>T (rs41528348, P=0.0118, MAF=0.10), is a control region (CR) sequence variant, and the non-coding regions are well known to contain regulatory elements affecting gene expression [24]. m.295C>T has been previously associated with low maximal oxygen uptake (VO2max) in response to a standardized aerobic exercise training program [100]. VO2max is a heritable trait, demonstrating a maternal effect [24]. VO2max is defined by the ability of the cardiorespiratory system to deliver oxygen to the exercising muscles, e.g., the heart rate and maximal cardiac output [7]. High VO2max (i.e., cardiorespiratory fitness) have been associated with a reduction in cardiovascular events and vice versa - low VO2max plays a role in increasing the risk of cardiovascular disease [54, 60, 93, 107]. Interestingly, an earlier study in UK Biobank [108], reported a significant association between m.295C>T and mean corpuscular hemoglobin (MCH) as well as mean corpuscular volume (MCV) (Fig.5). Both traits have been shown to increase with training [10]. In line with these earlier reports, our results show that m.295C>T was more frequent (OR=1.05; 95% CI 1.02-1.09, P=0.0118) in HARD cases, thus, potentially conferring a decreased cardiorespiratory fitness/exercise capacity and increased CAD risk. Of note, m.295C>T is also tagging macro-haplogroup J, and it has been previously shown that VO2max is lower in J than in non-J macro-haplogroup individuals [72] and excellence in endurance performance was less frequent among macro-haplogroup J individuals [52].

**Figure 5.**
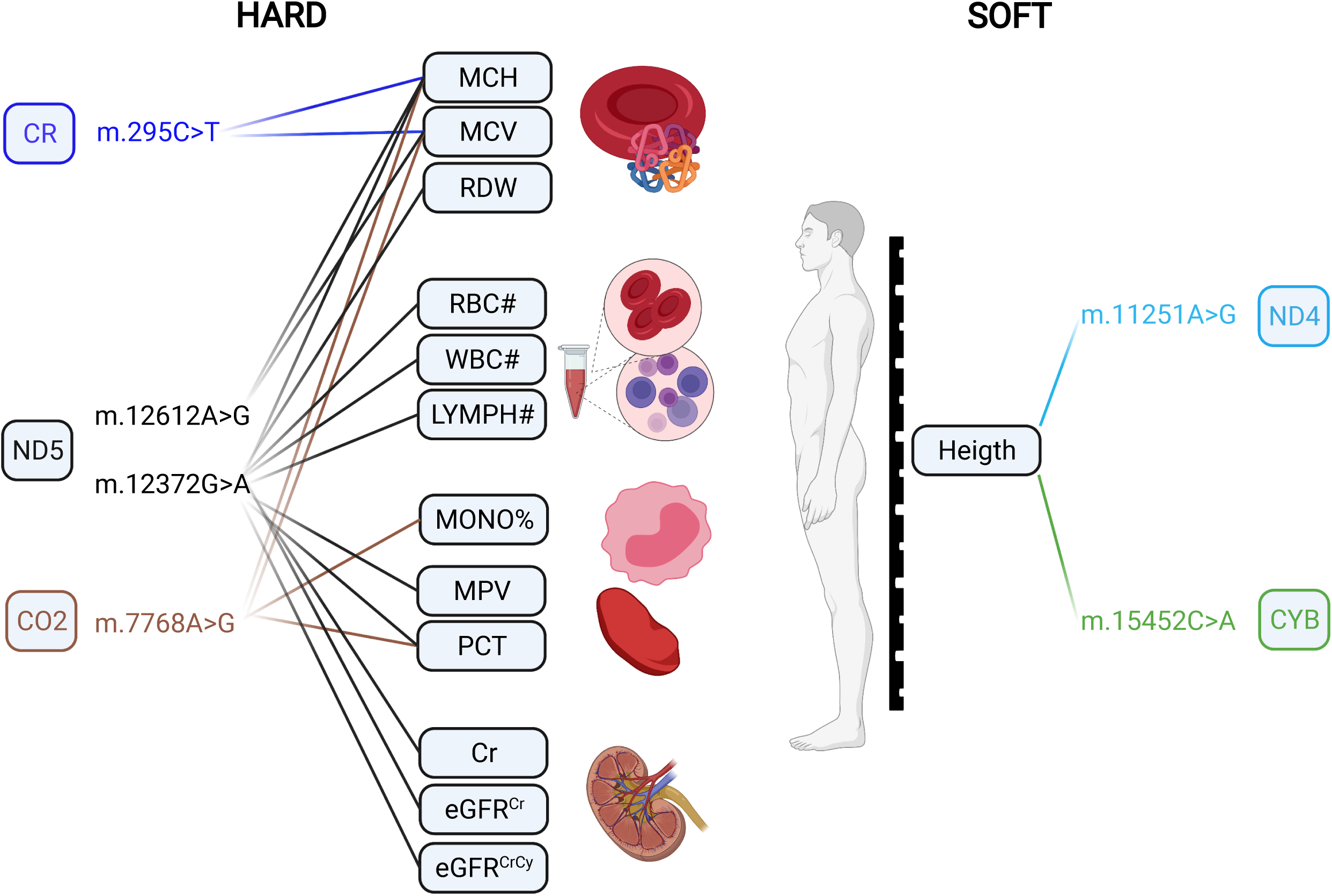
Previous findings for HARD and SOFT CAD-MT-SNV associations in UK Biobank by ref. by ref. [108]. MCH: Mean corpuscular hemoglobin; MCV: Mean corpuscular volume; RDW: Red blood cell distribution width; RBC#: Red blood cell count; WBC#: White blood cell count; LYMPH#: Lymphocyte count; MONO%: Monocyte percentage of white cells; MCV: Mean corpuscular volume; PCT: Plateletcrit; Cr: Creatinine; eGFRCr: Estimated glomerular filtration rate creatinine; eGFRCrCy: Estimated glomerular filtration rate creatinine and cystatin C. *Created with BioRender.com*.

m.12612A>G (rs28359172, P=0.0158, MAF=0.10) is a synonymous (V92V) sequence variant in the subunit 5 of NADH dehydrogenase (ND5), also tagging macro-haplogroup J and demonstrating significant associations with MCH and MCV (Fig.5) in UK Biobank [108]. In our study, it was also more frequent (OR=1.05; 95% CI 1.02-1.08, P=0.0158) in HARD cases. Therefore, it is reasonable to assume that m.295C>T also may be related to a decreased cardiorespiratory fitness/exercise ability and increased CAD risk. m.12372G>A (rs2853499, P=0.0059, MAF=0.22) is a synonymous (L12L) sequence variant in the ND5 gene, found more often in controls vs. HARD CAD cases (OR=0.97; 95% CI 0.95-0.99) and tagging macro-haplogroup U, members of which have been shown to represented higher brain pH than others [83]. m.11467A>G (rs2853493, P=0.0065, MAF=0.22) represents another synonymous (L236L) sequence variant also tagging macro-haplogroup U and more frequently found in controls vs. HARD CAD cases (OR=0.97; 95% CI 0.95-1.00), however, it is located in the subunit 4 of NADH dehydrogenase (ND4). m.11467A>G (rs2853493) has also been also associated with altered brain pH [83]. Higher brain pH has been shown to correlate with a lower brain activity and conferring protection against psychiatric disorders [41] and attention deficit/hyperactivity [15]. This protective role of macro-haplogroup U could be partially explained by reduced load of harmful reactions [15], as pH is known to play a role in mitochondrial ROS generation [90] and cybrids of macro-haplogroup U have been demonstrated to display lower inner mitochondrial membrane potential and mitochondrial protein synthesis and cell growth capacity [39]. Moreover, macro-haplogroup U has been associated with increased longevity that may be at least partly explained by its lower ROS production rate [17]. In our study, m.12372G>A (rs2853499) was found more frequently in controls, suggesting potential favorable differences in pH and ROS production in these subjects. Interestingly, in addition to associations with MCH and MCV, m.12372G>A displayed significant (P< 1e - 5) associations with eight additional blood cell and kidney-related traits in UK Biobank (Fig.5) [108]. Endurance time during exercise has been related to pre-exercise blood pH and demonstrated to increase with increasing pH [47]. m.7768A>G (rs41534044, P=0.0185, MAF=0.04) represents another synonymous (M61M) sequence variant located in the subunit 2 of mitochondrial encoded cytochrome c oxidase (CO2), more frequently found in controls vs. HARD CAD cases (OR=0.91; 95% CI 0.86-0.96) and displaying significant (P< 1e - 5) associations with several phenotypic traits in UK Biobank [108], including MCH and MCV, as well as the Monocyte percentage of white cells (MONO%) and Plateletcrit (PCT)(Fig.5).

In SOFT cases, four MT-SNVs survived multiple testing correction (at FDR<5%; Table 3 and Fig.3), all potentially conferring increased CAD risk: m.10400C>T, m.11251A>G, m.15452C>A and 15301G>A. m.11251A>G (rs869096886, P=0.0011, MAF=0.20) represents a synonymous sequence variant in ND4 gene and m.15452C>A (rs193302994, P=0.0017, MAF=0.20) is a non-synonymous (Leu → Ile) sequence variant in CYB gene, both were found more frequently in SOFT CAD cases vs. controls (OR=1.03; 95% CI 1.01-1.05). m.11251A>G (rs869096886) and m.15452C>A (rs193302994) are tagging macro-haplogroups J and T, potentially related to a decreased cardiorespiratory fitness/exercise capacity [52, 72] and thus increased CAD risk, as discussed above. Moreover, both MT-SNVs displayed significant (P< 1e - 5) associations with body height in UK Biobank (Fig.5) [108]. In line with this, we observed that the average body height of both male and female CAD cases was ∼2.00 cm lower compared to controls (Table 1). Previous studies have demonstrated that shorter body height is related to an increased CAD risk [78], which could be partly explained by altered lipid profile.

Gene-based tests revealed that in both, HARD and SOFT cases, CO2 displayed gene-based association at nominal significance (P<0.05, Table 4). The CO2 gene encodes for the second subunit of cytochrome c oxidase (COX, complex IV). Dysfunction of COX has been previously associated with mitochondrial oxidative stress, obesity and T2D [43]. CYB and ND3 were also associated (nominal P<0.05) with SOFT CAD phenotype (Table 4). Somatic variations in CYB have been previously related to hypertrophic cardiomyopathy (one of its clinical manifestations being angina) and exercise intolerance [40]. Recently, a large gene-based meta-analysis of mitochondrial genes with several metabolic traits identified ND3 associated with BMI (P<1e-03) [57].

All haplogroups demonstrating significant (at FDR<5%) associations in our study (M45a, G2b1a2, U2b2 with both HARD/SOFT, M57b1 and L2c (under-represented) with HARD and M3a with SOFT CAD phenotypes; Table 5 and Supplementary Table 4) were with a frequency <1%, while other studies have considered only haplogroups with a frequency ≥5% [26]. Low counts in the less common haplogroups may lead to a false-positive result [86]. Although, this should be addressed by performing multiple testing correction, grouping the less frequent haplogroups may be another approach to tackle this [86]. Hence, we also assigned individuals to one of the major European haplogroups (Fig.4) for comparison. As a result, we observed that 43.28% of the individuals belonged to the macro-haplogroup H, 13.70% to the macro-haplogroup U, 10.70% to the macro-haplogroup J and 9.52% to the macro-haplogroup T (Fig.4), in line with earlier reports in other European populations [2]. Overall, the frequencies of the major European mitochondrial haplogroups did not differ significantly (at FDR<5%) between CAD patients and control subjects (Fig.4). Only the frequency of haplogroup I was significantly (nominal P<0.05) higher in patients with HARD CAD phenotype vs. controls (3.35% vs. 3.08%, OR=1.09) and the macro-haplogroup R was significantly (nominal P<0.001 and P<0.01) higher in patients with HARD and SOFT CAD phenotype vs. controls (0.26% and 0.23% vs. 0.16%, OR=1.70 and OR=1.49, respectively) (Fig.4). A recent study [108] identified a significant effect of macro-haplogroup I on the MCH levels in UK Biobank. As previously discussed, this trait is known to increase with training [10], thus, again being potentially related to cardiorespiratory fitness/exercise capacity and thus CAD risk. Of note, however, we were able to assign most samples reliably into haplogroups as the mtDNA haplogroups were deduced from genotyping arrays with limited numbers of (high quality) SNVs being profiled, hence the quality score for haplogroup assignment ranged from 0.50 to 0.86, with a median of 0.68. Hence, we were not able to exclude samples with quality scores for haplogroup assignment <0.8 (as in ref. [36]). Moreover, we performed Fisher’s exact test that did not allow us to adjust for co-variates, hence it is possible that known and unknown potential confounding factors might have influenced these results. Though, in which cases to adjust for which covariates and whether it will increase or decrease the study power and/or bias, is still a matter of intense debate [5, 99].

Several other limitations should also be acknowledged. It is well-known that exceptionally large cohorts are required to reliably associate genetic variations with complex traits 86. The power for detecting causal MT-SNVs and haplogroups has been compared with that in the nuclear genome given equal effect sizes, estimating that the sample size needed for the mitochondrial studies would be roughly the same as that needed for the nuclear genome studies 73. Previous power calculations for ischemic stroke (assuming an additive model) [3] revealed a maximum power of 73% to detect SNVs with OR=1.4 and MAF=0.30, while for SNVs conferring OR=1.20 and MAF=0.20, the study power dropped to 4.6% and further to 0.001% for OR=1.10 and MAF=0.10. This study concluded that “prohibitively large sample sizes” would be needed to achieve sufficient power to detect individual MT DNA variants [3]. In line with this, we observe that in HARD CAD cases, where N=20,405, no MT-SNVs survived multiple testing correction, while when increasing N to 34,782 in SOFT CAD cases, four MT-SNVs survived multiple testing correction (FDR<5%). Hence, even larger sample sizes may be needed to reliably associate MT SNVs and haplogroups with CAD.

In addition to study individuals, also the number of MT-SNVs studied was limited. In UK Biobank [13], genotyping was performed using Affymetrix UK biobank Axiom (450,000 samples) and Affymetrix UK BiLEVE Axiom (50,000 samples) arrays, which included 265 genotyped MT DNA variants. After quality control procedures (Fig.1), 243 MT-SNVs remained for further analyses, 111 of those where common or low-frequency (MAF>0.01) and could be used for single marker association analyses. However, this is clearly not a representative set of MT-SNVs and, as previously recognized, some regions may be not well-covered, such as the hypervariable regions [32, 34]. Clearly, whole genome sequencing or targeted-sequencing of MT-DNA, considering their ability to achieve a deep genome coverage, would allow the identification many more MT-SNV (especially the low-frequency/rare variants; MAF<0.01), also improving the detection of haplogroups and allowing to investigate heteroplasmy, a phenomenon characteristic to MT DNA [18, 92].

Heteroplasmy denotes the coexistence of MT DNA genomes with wild-type inherited SNVs and somatic variants in varying ratios, which are dynamically determined and display patterns of cell and tissue specificity, and may differ even within the same mitochondrion [92], determining the clinical presentation of disease phenotypes [32, 33, 81]. In this study, we were limited to genotype calls from arrays, which are restricted in terms of minor alleles and do not allow to capture heteroplasmy [32, 33]. Moreover, MT DNA content was assessed only in blood cells, whereas previous studies have identified additional six vascular and metabolic tissues relevant to CAD [35, 94]. Therefore, whole genome sequencing/targeted-sequencing of MT-DNA, across several vascular and metabolic tissues relevant to CAD may be necessary [35, 94, 101] in order to characterize the full landscape of mitochondrial genetic variations and their potential contribution to these complex disease phenotypes. Especially, considering that the energy requirements and thus sensitivity to the changes in mitochondrial function differ for different cells and tissues and hence may be important in determining the phenotypic effect of MT SNVs [11].

We also do not consider mitochondrial DNA copy number (MT DNA-CN), representing the number of mitochondria per cell and the number of MT DNA per mitochondrion [6, 19]. Each mitochondrion contains multiple copies of MT DNA and different cells and tissues contain different numbers (up to 7,000) of mitochondria, again displaying patterns of cell and tissue variability [6, 19]. MT DNA-CN is believed to serve as an indirect biomarker that would capture the underlying mitochondrial activity and function, such as energy production capacity and metabolic characteristics, thus possibly playing a causative role in health and disease [19]. Decreased MT DNA-CN has been previously associated with an increased risk of developing cardiovascular disease (CVD) outcomes [6]. More recently, similar analyses in the UK Biobank demonstrated a possible causal role of lower MT DNA-CN on higher CAD risk [67]. In an even larger cohort (of 408,361 individuals from TOPMed and UK Biobank), a decline in MT DNA-CN was observed in elderly individuals (>65 years) and lower MT DNA-CN levels also demonstrated an age-independent associations with hypertension, hyperlipidemia, T2D and obesity, i.e., the well-known CAD risk factors [64]. However, none of these studies compared the MT DNA-CN levels between HARD vs. SOFT CAD phenotypes, which could be a subject of future studies. However, considering that MT DNA-CN varies greatly across cell and tissue types, again profiling of several vascular and metabolic tissues relevant to CAD may be necessary for such investigations [35, 94, 101].

Yet another important aspect not considered here is the nuclear genome, considering the co-evolution of mitochondria and eukaryotic cells [74]. The mitochondrial genome encodes only 37 genes, mainly components of the OXPHOS machinery, whereas the remaining ∼1000-1500 mitochondrial proteins are all encoded by the nuclear genome [42]. The importance of common genetic variation in the nuclear genome regulating MT heteroplasmy and DNA-CN is an active area of research [19, 38, 46, 75]. Moreover, genetic variants in nuclear genes could lead to oxidative disorders or modulate the mitochondrial variants [81] and mild nuclear gene variants could potentially become clinically relevant when combined with an incompatible MT DNA [105]. Additive interactions (epistasis) between mitochondrial variants in MT-ND2 gene and nuclear variants in genes responsible for mitochondrial replication and transcription have been demonstrated to influence the BMI and obesity phenotype [66]. Similarly, our previous investigations have demonstrated the role of nuclear encoded mitochondria imported genes in coordinating the response to hypercholesterolemia and atherosclerotic lesion expansion, and foam cell formation [102]. Hence, further analyses also considering these additional variations will be required.

Finally, similar to SNVs in nuclear genome, even if (mitochondria) genome-wide significant associations with HARD/SOFT CAD phenotypes would be identified, their functional consequences would need to be determined in the CAD relevant tissues [35, 94]. Currently, functional studies for MT-SNVs are not readily available, however, several novel experimental animal models (e.g. mice strains displaying DNA haplogroups similar to those observed in humans) may be available in the near future, allowing to investigate/define the potential causality of the relationship between inherited “natural” non-pathogenic MT-SNVs and potential alterations in mitochondrial function (e.g. oxygen consumption and oxidant production, cellular ATP levels) and their relation to alterations in cardiovascular function and CAD risk [11, 74, 81].

In conclusion, we found only spurious MT-SNV, gene and haplogorup associations with HARD and SOFT CAD phenotypes and conclude that whole genome sequencing/targeted-sequencing of MT-DNA, across several vascular and metabolic tissues relevant to CAD in even larger study cohorts (N>50,000), followed by functional studies in animal models, may be necessary to conclusively determine the role of MT-SNVs, genes and haplogorup in modulating the risk of CAD. Therefore, whole genome sequencing/targeted-sequencing of MT-DNA, across several vascular and metabolic tissues relevant to CAD may be necessary [35, 94, 101] in order to characterize the full landscape of mitochondrial genetic variations and their potential contribution to these complex disease phenotypes. Especially, considering that the energy requirements and thus sensitivity to the changes in mitochondrial function differ for different cells and tissues and hence may be important in determining the phenotypic effect of MT SNVs [11].

## Data Availability

All data produced will be submitted to the UK Biobank Resource (https://www.ukbiobank.ac.uk/) after successful publishing of this manuscript.

## Acknowledgments and Sources of Funding

This research was funded by the Latvian Council of Science, project, “Using Machine Learning to Model the Complex Interplay Between Diet, Genetic Factors and Mitochondria in Coronary Artery Disease”, project No. lzp-2020/2-0338. The present study has been conducted using the UK Biobank Resource (application ID 61684) that is available to researchers.

